# Place and causes of acute cardiovascular mortality during the COVID19 pandemic: retrospective cohort study of 580,972 deaths in England and Wales, 2014 to 2020

**DOI:** 10.1101/2020.07.14.20153734

**Authors:** Jianhua Wu, Mamas A Mamas, Mohamed Mohamed, Chun Shing Kwok, Chris Roebuck, Ben Humberstone, Tom Denwood, Tom Luescher, Mark A de Belder, John E Deanfield, Chris P Gale

## Abstract

**Importance:** The COVID-19 pandemic has resulted in a decline in admissions with cardiovascular (CV) emergencies. The fatal consequences of this are unknown.

**Objectives:** To describe the place and causes of acute CV death during the COVID-19 pandemic.

**Design:** Retrospective nationwide cohort.

**Setting:** England and Wales.

**Participants:** All adult (age ≥18 years) acute CV deaths (n=580,972) between 1^st^ January 2014 and 2^nd^ June 2020.

**Exposure:** The COVID-19 pandemic (defined as from the onset of the first COVID-19 death in England on 2^nd^ March 2020).

**Main outcomes:** Place (hospital, care home, home) and acute CV events directly contributing to death as stated on the first part of the Medical Certificate of Cause of Death.

**Results:** After 2^nd^ March 2020, there were 22,820 acute CV deaths of which 5.7% related to COVID-19, and an excess acute CV mortality of 1752 (+8%) compared with the expected daily deaths in the same period. Deaths in the community accounted for nearly half of all deaths during this period. Care homes had the greatest increase in excess acute CV deaths (1065, +40%), followed by deaths at home (1728, +34%) and in hospital (57, +0%). The most frequent cause of acute CV death during this period was stroke (8,290, 36.3%), followed by acute coronary syndrome (ACS) (5,532, 24.2%), heart failure (5,280, 23.1%), pulmonary embolism (2,067, 9.1%) and cardiac arrest (1,037, 4.5%). Deep vein thrombosis had the greatest increase in cause of excess acute CV death (18, +25%), followed pulmonary embolism (340, +19%) and stroke (782, +10%). The greatest cause of excess CV death in care homes was stroke (700, +48%), compared with cardiac arrest (80, +56%) at home, and pulmonary embolism (126, +14%) and cardiogenic shock (41, +14%) in hospital.

**Conclusions and relevance:** The COVID-19 pandemic has resulted in an inflation in acute CV deaths above that expected for the time of year, nearly half of which occurred in the community. The most common cause of acute CV death was stroke followed by acute coronary syndrome and heart failure. This is key information to optimise messaging to the public and enable health resource planning.

## Introduction

Cardiovascular disease (CVD) is one of the most prevalent underlying condition associated with increased mortality from COVID-19 infection.^1-5^ Yet, we and others have shown a substantial reduction in presentations to hospitals with acute cardiovascular (CV) conditions including acute coronary syndrome, heart failure, cardiac arrhythmia and stroke during the pandemic.^6-9^. This would be expected to result in a much higher number of deaths, unless there has been an actual decrease in the incidence of these acute conditions. The detailed impact on mortality from acute CVD has, however, not been studied at national level.

We now report, with high temporal resolution, CV specific mortality during COVID-19 in England and Wales. In particular, we have evaluated the location of CV deaths (e.g. hospitals, home or care homes), their relation to COVID-19 infection and the specific CV fatal events that contributed directly to death. This information is vital for the understanding of healthcare policy during the pandemic and to assist Governments around the world reorganise healthcare services.

## Methods

### Data and deaths

The analytical cohort included all certified and registered deaths in England and Wales ≥18 years of age, between 1^st^ January 2014 and 2^nd^ June 2020 recorded in the Civil Registration Deaths Data of the Office for National Statistics (ONS) of England and Wales.^10^

### Acute CV death

The primary analysis was based upon any of the ICD-10 codes corresponding to the immediate cause of death and contributed causes registered, and the doctor who attended the deceased during their last illness completes a medical certificate of cause of death (MCCD) within 5 days unless there is to be a coroner’s post-mortem or an inquest. Cardiovascular events directly leading to death (herein called acute CV deaths) were categorised as acute coronary syndrome (ST-elevation myocardial infarction (STEMI), non-STEMI (NSTEMI), type 2 myocardial infarction, re-infarction) abbreviated as ACS, heart failure, cardiac arrest, ventricular tachycardia and/or ventricular fibrillation (VT and VF), stroke (acute ischaemic stroke, acute haemorrhagic stroke, other non-cerebral strokes, unspecified stroke), cardiogenic shock, pulmonary embolism, deep venous thrombosis, aortic disease (aortic aneurysm rupture and aortic dissection) and infective endocarditis (Supplement table 1). ICD-10 codes ‘U071’ (confirmed) and ‘U072’ (suspected) were used to identify whether a death was related to COVID-19 infection on any part of the MCCD.

### Statistical Analyses

Baseline characteristics were described using numbers and percentages for categorical data. Data were stratified by COVID-19 status (suspected or confirmed, not infected), age band (<50, 50-59, 60-69, 70-79, 80+ years)), sex, place of death (home, hospital, care home or hospice). The number of daily deaths was presented using a 7-day simple moving average (the mean number of daily deaths for that day and the preceding 6 days) from 1^st^ February 2020 up to and including 2^nd^ June 2020, adjusted for seasonality.

The expected daily deaths from 1^st^ February 2020 up to and including 2^nd^ June 2020 were estimated using Farrington surveillance algorithm for daily historical data between 2014 and 2020.^11^ The algorithm used overdispersed Poisson generalised linear models with spline terms to model trends in counts of daily death, accounting for seasonality. The number of non-COVID-19 CV deaths each day from 1^st^ February 2020 were subtracted from the estimated expected daily deaths in the same time period to create a zero historical baseline. Deaths above this baseline maybe interpreted as excess mortality, which were calculated as the difference between the observed daily deaths and the expected daily deaths. Negative values, where the observed deaths fell below the expected deaths, were set to zero. The rate of excess deaths was derived from dividing excess mortality by the sum of the expected deaths between 2^nd^ March 2020 and 2^nd^ June 2020.

For the categories of acute CV death, the ICD-10 code on the MCCD was counted only once per deceased. Thus, the overall rate of acute CV death represents the number of people with a direct CV cause of death. Given that, people may have more than one of the pre-defined CV events leading to death, analyses for each of the pre-defined CV categories represents the number of events (not people) per category. For the purposes of this investigation, CVD that contributed, but did not directly lead to death were excluded from the analyses. All tests were two-sided and statistical significance considered as *P*<0.05. Statistical analyses were performed in R version 4.0.0.

## Results

Between 1^st^ January 2014 and 2^nd^ June 2020, there were 3,401,270 deaths from all-causes among adults. The proportion of deaths increased with increasing age band and there were 1,728,591 (50.8%) in women (Table 1). People dying from any of the directly contributing CV categories accounted for 58,0972 (17.1%) of all deaths, of which 6.1% had at least two of the pre-defined CV categories that directly contributed to death. Most deaths occurred in hospital (62.6%) followed by home (23.7%) and at care home (13.6%).

**Table 1.** Acute CV deaths before and after 2^nd^March 2020, by COVID-19 status.

|  | Acute CV deaths before 2<sup>nd</sup> March 2020 | Non-COVID-19 acute CV deaths after 2<sup>nd</sup> March 2020 | COVID-19 acute CV deaths after 2<sup>nd</sup> March 2020 | Acute CV deaths after 2<sup>nd</sup> March 2020 |
| --- | --- | --- | --- | --- |
| **Total** | n = 558,152 | n = 21,513 | n = 1,307 | n = 22,820 |
| **Sex** |  |  |  |  |
| Men | 288,770 (51.7) | 11,440 (53.2) | 768 (58.8) | 12,208 (53.5) |
| Women | 269,382 (48.3) | 10,073 (46.8) | 539 (41.2) | 10,612 (46.5) |
| **Age category (years)** |  |  |  |  |
| 18-49 | 20,182 ( 3.6) | 850 ( 4.0) | 51 ( 3.9) | 901 ( 3.9) |
| 50-59 | 32,667 ( 5.9) | 1,419 ( 6.6) | 116 ( 8.9) | 1,535 ( 6.7) |
| 60-69 | 64,743 (11.6) | 2,644 (12.3) | 185 (14.2) | 2,829 (12.4) |
| 70-79 | 128,845 (23.1) | 5,252 (24.4) | 335 (25.6) | 5,587 (24.5) |
| 80+ | 311,715 (55.8) | 11,348 (52.7) | 620 (47.4) | 11,968 (52.4) |
| **Region** |  |  |  |  |
| North East | 24,113 ( 5.1) | 1,074 ( 5.0) | 55 ( 4.2) | 1,129 ( 4.9) |
| North West | 65,429 (13.7) | 2,875 (13.4) | 166 (12.7) | 3,041 (13.3) |
| Yorkshire and The Humber | 48,026 (10.1) | 2,121 ( 9.9) | 119 ( 9.1) | 2,240 ( 9.8) |
| East Midlands | 36,534 ( 7.7) | 1,681 ( 7.8) | 77 ( 5.9) | 1,758 ( 7.7) |
| West Midlands | 51,883 (10.9) | 2,399 (11.2) | 178 (13.6) | 2,577 (11.3) |
| East of England | 49,023 (10.3) | 2,214 (10.3) | 110 ( 8.4) | 2,324 (10.2) |
| London | 50,823 (10.7) | 2,277 (10.6) | 253 (19.4) | 2,530 (11.1) |
| South East | 71,504 (15.0) | 3,226 (15.0) | 204 (15.6) | 3,430 (15.0) |
| South West | 49,513 (10.4) | 2,373 (11.0) | 75 ( 5.7) | 2,448 (10.7) |
| Wales | 29,306 ( 6.2) | 1,273 ( 5.9) | 69 ( 5.3) | 1,342 ( 5.9) |
| **Place of death\*** |  |  |  |  |
| Care home and hospice | 73,967 (13.5) | 3,542 (16.8) | 148 (11.4) | 3,690 (16.5) |
| Home | 128,370 (23.5) | 6,683 (31.8) | 93 ( 7.1) | 6,776 (30.4) |
| Hospital | 345,028 (63.0) | 10,799 (51.4) | 1,061 (81.5) | 11,860 (53.1) |
| **Underlying acute CV cause of deaths\*\*** |  |  |  |  |
| Stroke | 199,730 (35.8) | 7,789 (36.2) | 501 (38.3) | 8,290 (36.3) |
| Acute coronary syndrome | 140,316 (25.1) | 5,256 (24.4) | 276 (21.1) | 5,532 (24.2) |
| Heart failure | 122,138 (21.9) | 5,048 (23.5) | 232 (17.8) | 5,280 (23.1) |
| Pulmonary embolism | 50,744 (9.1) | 1,844 (8.6) | 223 (17.1) | 2,067 (9.1) |
| Cardiac arrest | 29,255 (5.2) | 954 (4.4) | 83 (6.4) | 1,037 (4.5) |
| Aortic diseases | 28,446 (5.1) | 888 (4.1) | 4 (0.3) | 892 (3.9) |
| Infective endocarditis | 13,499 (2.4) | 648 (3.0) | 28 (2.1) | 676 (3.0) |
| Cardiogenic shock | 6,596 (1.2) | 205 (1.0) | 13 (1.0) | 218 (1.0) |
| VT and VF | 2,356 (0.4) | 73 (0.3) | 3 (0.2) | 76 (0.3) |
| Deep vein thrombosis | 480 (0.1) | 74 (0.3) | 2 (0.2) | 76 (0.3) |

CV: cardiovascular; VT: ventricular tachycardia; VF: ventricular fibrillation
\*The numbers do not add up to the total deaths due to missingness (1.9%).
\*\*Listed CV related conditions may not add up to 100 percent because some deaths may have multiple CV related conditions.

Acute CV deaths before 2^nd^ March 2020 Of the 558,152 acute CV deaths between 1^st^ January 2014 and 1^st^ March 2020, 48.3% were among women (Table 1). The most frequent place of death was hospital and most frequent cause of death was stroke, which accounted for 199,730 (35.8%) of the acute CV deaths. The South East of England had the greatest number of acute CV deaths.

Acute CV deaths after 2^nd^ March 2020 Following the first UK death from COVID-19, there were 22,820 acute CV deaths of which 5.7% related to COVID-19 (7.8% suspected; 92.2% confirmed), and an excess acute CV mortality of 1752 (+8%) compared with the expected historical average in the same time period of the year. COVID-19 deaths accounted for 1,307 (74.6%) of all excess deaths after this date (Figure 1, Table 2). Qualitatively, excess acute CV mortality began in late March 2020 and peaked in early April 2020. Whist hospital remained the most frequent place of acute CV death, there were proportionally fewer deaths in hospital (53.1% vs 63.0%) and more at home (30.4% vs 23.5%) and in care homes (16.5% vs 13.5%) (Table 1). Moreover, care homes had the greatest increase in excess acute CV deaths (+40%), followed by deaths at home (+34%) and there was no excess acute CV deaths in hospital (Table 2). The number of excess acute CV deaths were higher more among men than women (1009, +9% vs. 773, +8%), and was highest in the age category 18-49 years (138, +17%) (Table 2). The most frequent cause of acute CV death during the COVID19 pandemic was stroke (36.3%), followed by acute coronary syndrome (24.2%), heart failure (23.1%), pulmonary embolism (9.1%) and cardiac arrest (4.5%) (Table 1). Moreover, deep vein thrombosis demonstrated the greatest increase in excess acute CV death (18, +25%), followed by pulmonary embolism (340, +19%) and stroke (782, +10%) (Figure 2, Table 2).

**Table 2.** COVID-19 related and total excess acute CV deaths.

|  | Excess acute CV deaths |  |
| --- | --- | --- |
|  | COVID-19 related | Total* |
| **Total** | 1,307 (+6%) | 1,752 (+8%) |
| **Sex** |  |  |
| Men | 768 (+7%) | 1,009 (+9%) |
| Women | 539 (+5%) | 773 (+8%) |
| **Age category (years)** |  |  |
| 18-49 | 51 (+7%) | 138 (+17%) |
| 50-59 | 116 (+8%) | 186 (+13%) |
| 60-69 | 185 (+7%) | 376 (+15%) |
| 70-79 | 335 (+7%) | 577 (+11%) |
| 80+ | 620 (+5%) | 651 (+6%) |
| **Region** |  |  |
| North East | 55 (+5%) | 54 (+5%) |
| North West | 166 (+5%) | 138 (+4%) |
| Yorkshire and The Humber | 119 (+5%) | 131 (+6%) |
| East Midlands | 77 (+4%) | 89 (+5%) |
| West Midlands | 178 (+8%) | 267 (+11%) |
| East of England | 110 (+5%) | 119 (+5%) |
| London | 253 (+11%) | 296 (+13%) |
| South East | 204 (+6%) | 157 (+5%) |
| South West | 75 (+3%) | 153 (+7%) |
| Wales | 69 (+5%) | 40 (+3%) |
| **Place of death\*\*** |  |  |
| Care home and hospice | 148 (+6%) | 1,065 (+40%) |
| Home | 93 (+2%) | 1,728 (+34%) |
| Hospital | 1,061 (+8%) | 57 (+0%) |
| **Underlying acute CV cause of deaths** |  |  |
| Stroke | 501 (+7%) | 782 (+10%) |
| Acute coronary syndrome | 276 (+5%) | 411 (+8%) |
| Heart failure | 232 (+5%) | 447 (+9%) |
| Pulmonary embolism | 223 (+13%) | 340 (+19%) |
| Cardiac arrest | 83 (+7%) | 23 (+2%) |
| Aortic diseases | 4 (+0%) | 8 (+1%) |
| Infective endocarditis | 28 (+4%) | 79 (+12%) |
| Cardiogenic shock | 13 (+4%) | 40 (+13%) |
| VT and VF | 3 (+2%) | 20 (+13%) |
| Deep vein thrombosis | 2 (+3%) | 18 (+25%) |

CV: cardiovascular; VT: ventricular tachycardia; VF: ventricular fibrillation
\*The excess death in subcategories may not add up to the total excess deaths due to estimation and rounding error when comparing to the respective historical average.
\*\*The excess death in place of death do not add up to the total excess deaths due to setting those daily deaths below the expected historical average to zeros in the respective subgroup

**Figure 1.**
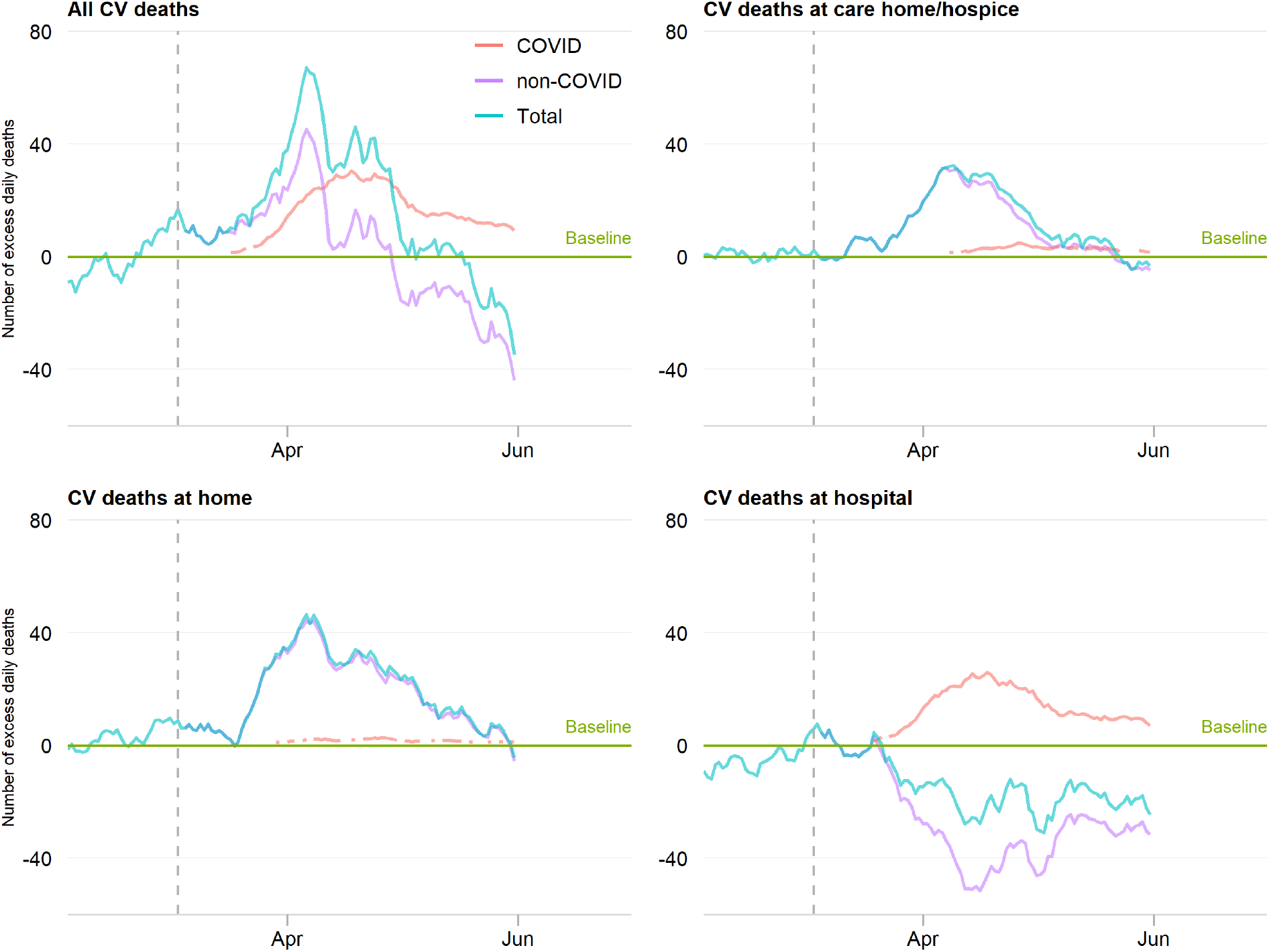
Time series of acute CV deaths, by place of death. The number of daily CV deaths is presented using a 7-day simple moving average (indicating the mean number of daily CV deaths for that day and the preceding 6 days) from 1^st^ February 2020 up to and including 2^nd^ June 2020, adjusted for seasonality. The number of non-COVID-19 excess CV deaths each day from 1^st^ February 2020 were subtracted from the expected daily death estimated using Farrington surveillance algorithm in the same time period. The green line is a zero historical baseline. The red line represents daily COVID-19 CV death from 2^nd^ March to 2^nd^ June 2020, the purple line represents excess daily non-COVID-19 CV death from 2^nd^ March to 2^nd^ June 2020 and the blue line represents the total excess daily CV death from 1^st^ February to 2^nd^ June 2020.

**Figure 2.**
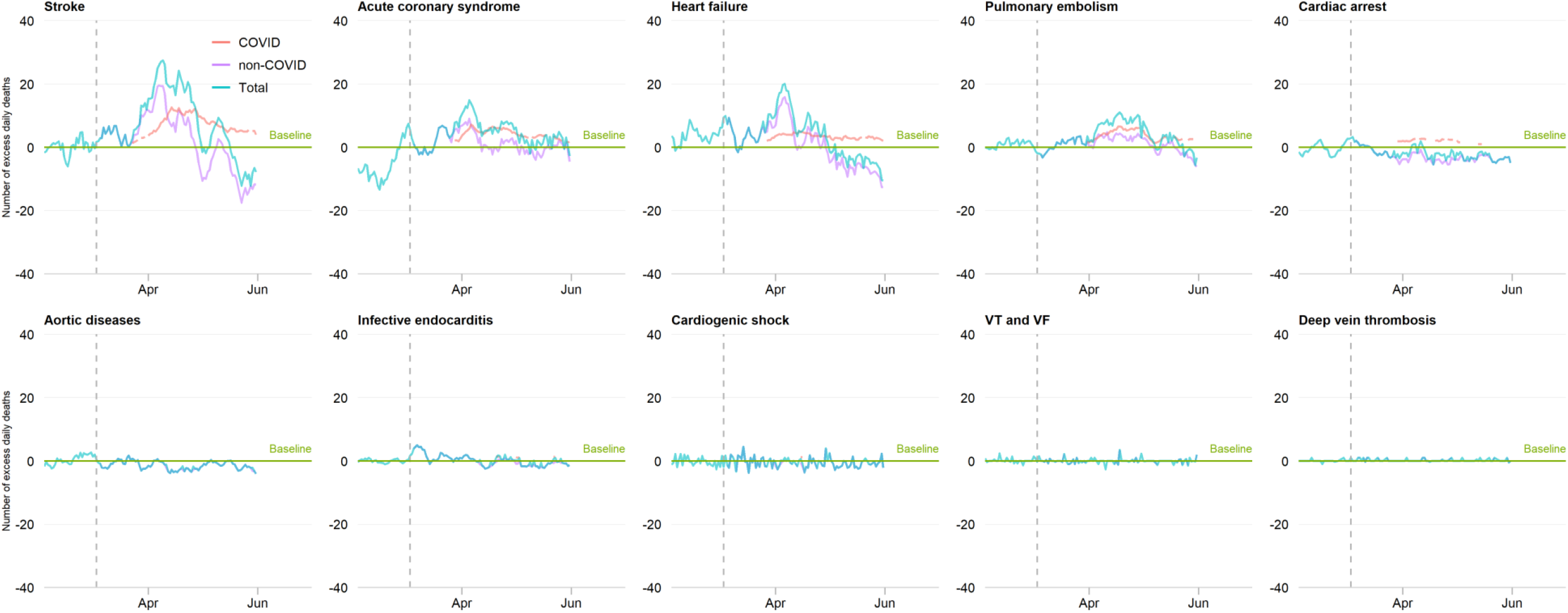
Time series of acute CV deaths by COVID-19, by cause of death. The number of daily CV deaths is presented using a 7-day simple moving average (indicating the mean number of daily CV deaths for that day and the preceding 6 days) from 1^st^ February 2020 up to and including 2^nd^ June 2020, adjusted for seasonality. The number of non-COVID-19 excess CV deaths each day from 1^st^ February 2020 were subtracted from the expected daily death estimated using Farrington surveillance algorithm in the same time period. The green line is a zero historical baseline. The red line represents daily COVID-19 CV death from 2^nd^ March to 2^nd^ June 2020, the purple line represents excess daily non-COVID-19 CV death from 2^nd^ March to 2^nd^ June 2020 and the blue line represents the total excess daily CV death from 1^st^ February to 2^nd^ June 2020.

COVID-19 CV deaths following 2^nd^ March 2020 Compared with deaths prior to 2^nd^ March 2020, COVID-19 CV deaths were more likely to occur in hospital (81.5% vs. 63.0%), much less at home (7.1% vs. 23.5%) and remained of similar proportions to non-COVID-19 CV deaths in care homes (13.5% vs. 11.4%). The rate of COVID-19 excess CV deaths was higher in hospitals than care homes (+8% vs. +6%), and less at home (+2%). Excess COVID-19 CV deaths occurred in similar proportions for men and women (+7% vs. +5%), and the rate of excess COVID-19 CV deaths was comparable across the age bands (Table 2). Pulmonary embolism had the greatest increase in cause of excess COVID-19 CV death (223, +13%), followed by stroke (501, +7%) and cardiac arrest (83, +7%) (Figure 2, Table 2).

Place and cause of death after 2^nd^ March 2020 The greatest inflation in excess CV death in care homes was for stroke (700, +48%), followed by acute coronary syndrome (116, +42%), and compared with cardiac arrest (80, +56%) and stroke (349, +47%) at home, and pulmonary embolism (126, +14%) and cardiogenic shock (41, +14%) in hospital (Figure 3, Table 3). For stroke, ACS, heart failure and cardiac arrest, the numbers of deaths in hospital were lower than the historical baseline (Figure 3).

**Table 3.** Excess acute CV deaths by cause and place of death.

|  | Care home and hospice |  | Home |  | Hospital |  |
| --- | --- | --- | --- | --- | --- | --- |
|  | COVID-19 related | Total | COVID-19 related | Total | COVID-19 related | Total* |
| Stroke | 82 (+6%) | 700 (+48%) | 16 (+2%) | 349 (+47%) | 398 (+7%) | 16 (+0%) |
| Acute coronary syndrome | 17 (+6%) | 116 (+42%) | 19 (+1%) | 621 (+42%) | 234 (+7%) | 25 (+1%) |
| Heart failure | 31 (+4%) | 218 (+31%) | 31 (+2%) | 531 (+31%) | 167 (+7%) | 32 (+1%) |
| Pulmonary embolism | 11 (+6%) | 38 (+22%) | 16 (+2%) | 198 (+31%) | 195 (+21%) | 126 (+14%) |
| Cardiac arrest | 5 (+5%) | 11 (+11%) | 13 (+9%) | 80 (+56%) | 64 (+6%) | 13 (+1%) |
| Aortic diseases | 0 (0%) | 0 (+19%) | 0 (0%) | 16 (+4%) | 4 (+1%) | 14 (+2%) |
| Infective endocarditis | 2 (+15%) | 4 (+29%) | 0 (0%) | 36 (+22%) | 24 (+6%) | 35 (+9%) |
| Cardiogenic shock | 0 (0%) | 0 (0%) | 0 (0%) | 0 (0%) | 13 (+4%) | 41 (+14%) |
| VT and VF | 0 (0%) | 0 (0%) | 0 (0%) | 1 (+3%) | 3 (+2%) | 14 (+9%) |
| Deep vein thrombosis | 0 (0%) | 0 (0%) | 0 (0%) | 1 (+4%) | 2 (+4%) | 5 (+9%) |

CV: cardiovascular; VT: ventricular tachycardia; VF: ventricular fibrillation
\*the positive excess rate in hospital was due to setting those daily deaths below the expected historical average to zeros.

**Figure 3.**
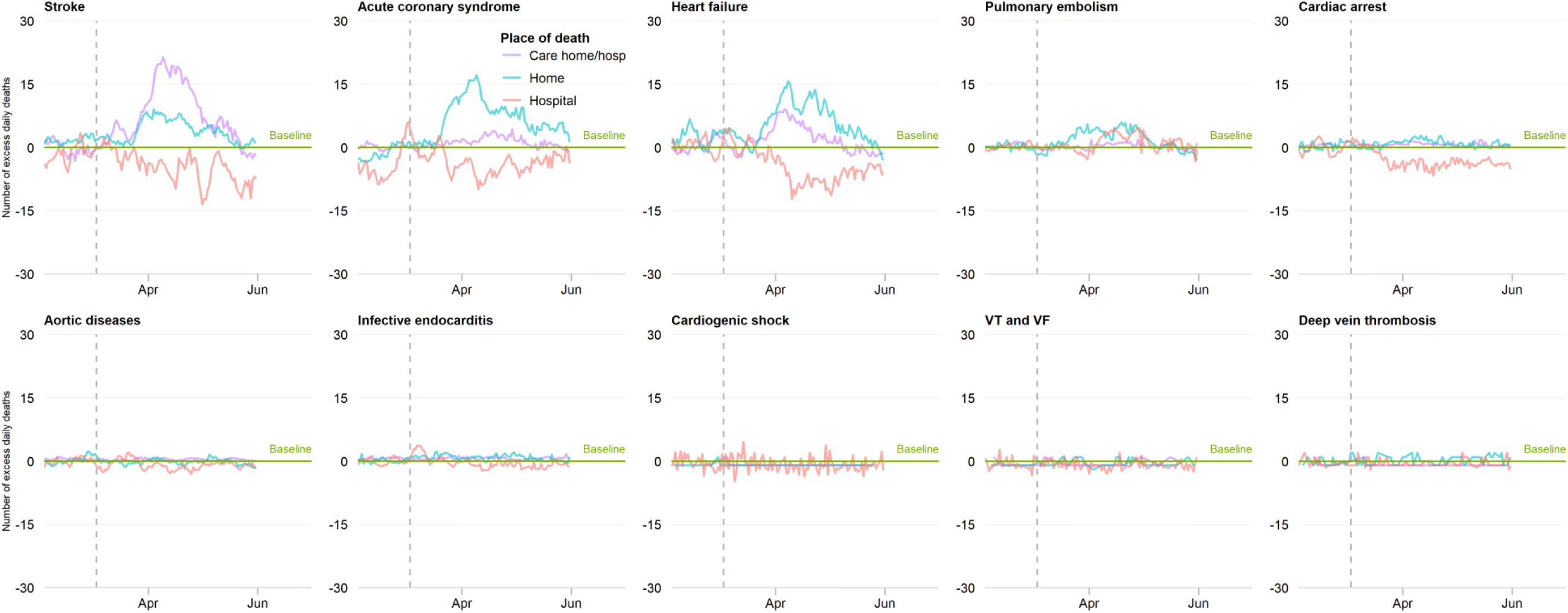
Time series of acute CV deaths by cause of death and place of death. The number of daily CV deaths is presented using a 7-day simple moving average (indicating the mean number of daily CV deaths for that day and the preceding 6 days) from 1^st^ February 2020 up to and including 2^nd^ June 2020, adjusted for seasonality. The number of non-COVID-19 excess CV deaths each day from 1^st^ February 2020 were subtracted from the expected daily death estimated using Farrington surveillance algorithm in the same time period. The green line is a zero historical baseline. The red line represents excess daily death at hospital, the purple line represents excess daily CV death at care home and hospice and the blue line represents excess daily CV death at home.

## Discussion

We show for the first time, in a nationwide complete analysis of all adult deaths, the extent, site and underlying causes of the increased acute CV mortality during the COVID-19 pandemic compared with previous years. This shows that the pandemic has resulted in an inflation in acute CV deaths above that expected for the time of year, nearly half of which occurred outside of the hospital setting, either at home or in care homes, and with care homes experiencing the greatest increase in excess acute CV deaths. The most common cause of acute CV death during the COVID19 pandemic was stroke followed by acute coronary syndrome and heart failure. This is key information to optimise messaging to the public, as well as for allocation of health resources and planning.

Numerous international studies have reported the decline in hospital presentations for a range of CV emergencies.^6-9^ To the best of our knowledge, this is the first study to show that this is associated with an adverse overall CV impact. Whilst stroke and acute coronary syndrome accounted for the vast majority of acute CV deaths, the number of deaths in hospital due to these conditions fell below that expected for the time of year and it increased in the community. This ‘displacement of death’, most likely, signifies that people either did not seek help or were not referred to hospital during the pandemic. Given the times series plots show that the excess in acute CV mortality began in late March 2020 and peaked in early April 2020, government directives at the time including the onset of the UK lockdown on 23^rd^ March 2020 could have accentuated the public response.

Care homes witnessed the greatest increase in excess acute CV deaths. Here, stroke, acute coronary syndrome, heart failure and pulmonary embolism were the commonest cause of acute CV death. This finding highlights the susceptibility of the elderly and co-morbid to the wider implications of COVID-19 crisis. That is, not only were care home residents prone to the respiratory effects of COVID-19 infection, but they will also have been exposed to the acute CV complications of COVID-19 and decisions not to go to hospital for fear of becoming infected. This situation will have been exacerbated by the discharge of unknowingly infected patients from hospitals to care homes early in the course of the pandemic, a lack of systematic antibody testing for the SARS-CoV-2 virus, the efficient person-to-person transmission of the virus and its propensity to death in the vulnerable.^1 12^

The major causes of acute CV death were different between hospital and community settings. In hospital, the greatest increase in excess acute CV deaths was for pulmonary embolism and cardiogenic shock, followed equally by ventricular tachycardia and ventricular fibrillation, deep vein thrombosis and infective endocarditis. Our earlier work using data from National Health Service hospitals in England found that during the COVID-19 pandemic, patients with acute myocardial infarction who did present to hospital delayed seeking help. The excess in deaths from cardiogenic shock and cardiac arrhythmias is, therefore, very likely the consequence of delayed presentation to hospital with acute coronary syndrome.

Complications of untreated acute myocardial infarction include cardiac arrest, arrhythmia, acute heart failure and stroke, many of which we found were recorded in excess early on in the pandemic. In line with previous findings,^13^ we also identified an increase in cardiac arrest – much more so at home than in hospital. Again, this signals the catastrophic impact of the delays to seeking help for acute CV conditions. In hospital, there was also an inflation of deaths from infective endocarditis and aortic dissection and rupture, indicating perhaps a more advanced (and for some, irreversible) stage of disease presentation during the pandemic.

Notwithstanding delays to seeking help, it is possible that COVID-19 is a critical factor in the pathophysiology of CV events. Infection with COVID-19 is associated with unrecognised venous and arterial thromboembolic complications and a COVID19 coagulopathy that confers an increased risk of death.^14-16^ Our study provides evidence for an increase in deaths above that expected for the time of year directly related to pulmonary embolism and deep vein thrombosis. Together, these findings lend support to the use of anticoagulation among higher risk people with COVID-19.^17^

Whilst previous reports have described an elevated risk of death among the elderly and people with CV disease during the COVID-19 pandemic, none have characterised the CV events directly leading to death or quantified the excess in acute CV mortality.^1 3 18^ To date, insights have been derived from small series of cases, regional or national death records data – each reporting elevated mortality rates, but none by the type and place of cardiovascular death.^1 2 19-22^ The unique strengths of this investigation include full population coverage of all adult deaths across places of death. Most previous reports have been confined to hospitals deaths and have not captured the full extent of the impact of the pandemic, including deaths outside of hospitals in people who may not have been tested for the disease.

Nonetheless, our study has limitations. During the COVID19 pandemic, emergency guidance enabled any doctor in the UK (not just the attending) to complete the MCCD, the duration of time over which the deceased was not seen before referral to the coroner was extended from 14 to 28 days, and causes of death could be “to the best of their knowledge and belief” without diagnostic proof, if appropriate and to avoid delay.^23^ This may have resulted in misclassification bias, with underreporting of the deaths directly due to CV disease in preference to COVID19 infection (which is a notifiable disease under the Health Protection (Notification) Regulations 2010) or respiratory disease. In fact, we found that MCCDs with COVID-19 certification less frequently contained details of acute CV events directly leading to death. Although the MCCD allows the detailing of the sequence of events directly leading to death, we found that after 2^nd^ March 2020 few (5.7%) had multiple acute CV events recorded, and therefore the categorisation of the acute CV events effectively represents per patient events. The lower proportion of deaths with COVID-19 at home and in care homes may represent the lack of access to community-based COVID19 testing. Equally, because there was no systematic testing of the UK populace for the presence the COVID-19, deaths associated with the infection may have been under estimated.^24^ This analysis will have excluded a small proportion of deaths under review by the Coroner, though typically these will have been unnatural in aetiology.

## Conclusion

To date, there is no whole-population, high temporal resolution information about CV-specific mortality during the COVID19 pandemic. Through the systematic classification of all adult deaths in England and Wales it has been possible to show that there has been an excess in acute CV mortality during the COVID-19 pandemic, seen greatest in care homes and which corresponds with the onset of public messaging and the substantial decline in admissions to hospital with acute CV emergencies

## Data Availability

The Secretary of State for Health and Social Care has issued a time limited Notice under Regulation 3(4) of the NHS (Control of Patient Information Regulations) 2002 (COPI) to share confidential patient information.

## Acknowledgments

We acknowledge the intellectual input of Professor Colin Baigent, University of Oxford.

JW had full access to all of the data in the study and takes responsibility for the accuracy of the data analysis. The Office for National Statistics provided NHS Digital with the mortality data and takes responsibility for the integrity of these data.

## Details of funding

JW and CPG are funded by the University of Leeds. MAM is funded by the University of Keele. The funding organizations for this study had no involvement in the design and conduct of the study; collection, management, analysis and interpretation of the data; preparation, review, or approval of the manuscript; or the decision to submit the manuscript for publication.

### Ethical approval

Ethical approval was not required as this study used fully anonymised routinely collected civil registration deaths data. The data analysis was conducted through remote access to NHS Digital Data Science Server.

### Data sharing

The Secretary of State for Health and Social Care has issued a time limited Notice under Regulation 3(4) of the NHS (Control of Patient Information Regulations) 2002 (COPI) to share confidential patient information. The study complies with the Declaration of Helsinki.

### Patient and Public Involvement statement

Patient and public were not involved because this study was to analyse routinely collected mortality data.

---

**Supplementary table 1.**
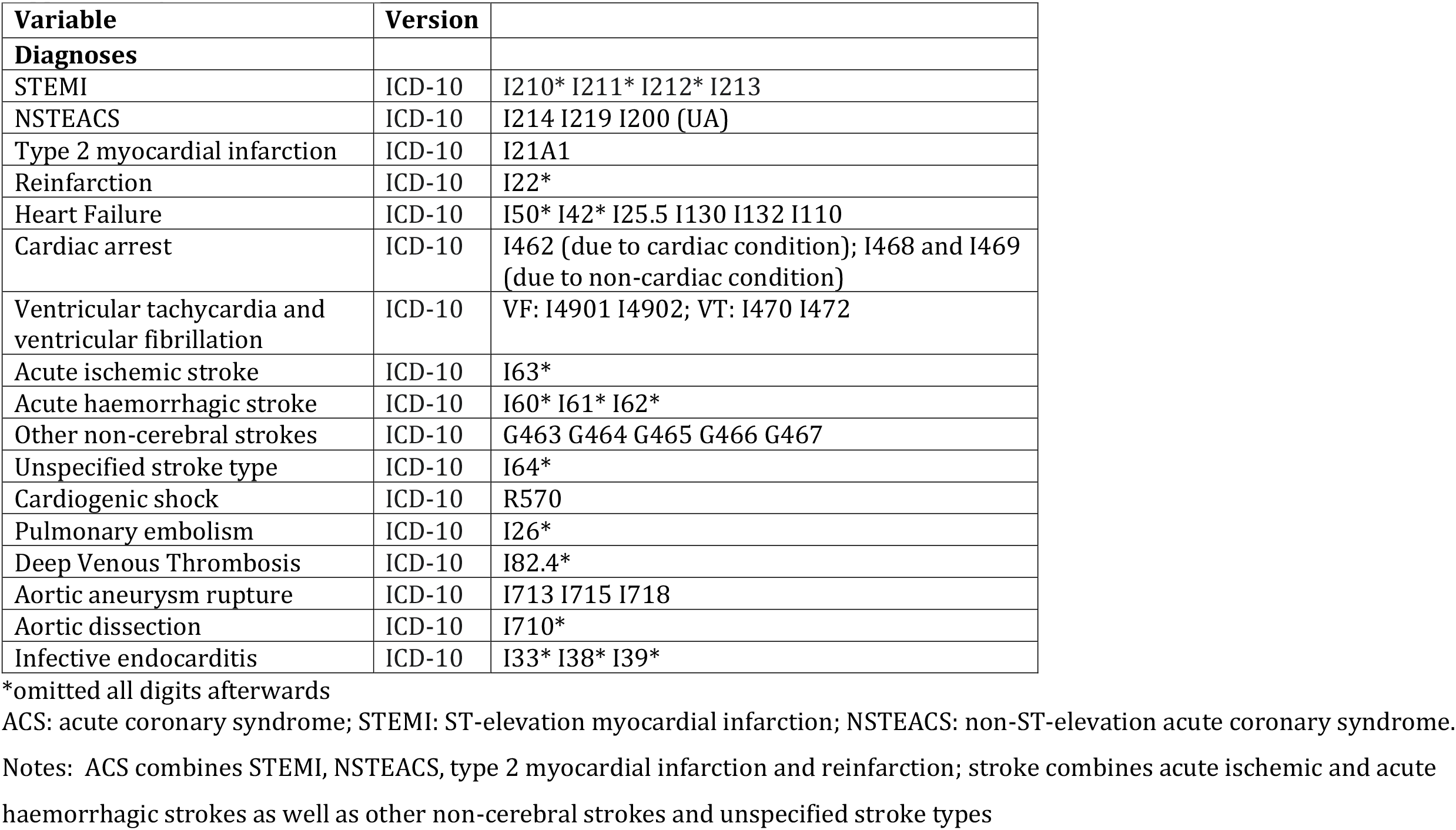
ICD-10 codes used to define acute cardiovascular causes of death

